# Mobility was a Significant Determinant of Reported COVID-19 Incidence During the Omicron Surge in the Most Populous U.S. Counties

**DOI:** 10.1101/2022.03.16.22272523

**Authors:** Jeffrey E. Harris

**Affiliations:** Massachusetts Institute of Technology, Cambridge MA 02139; Eisner Health, Los Angeles CA 90015

## Abstract

**Background:** Significant immune escape by the Omicron variant, along with the emergence of widespread worry fatigue, have called into question the robustness of the previously observed relation between population mobility and COVID-19 incidence.

**Methods:** We employed principal component analysis to construct a one-dimensional summary indicator of six Google mobility categories. We related this mobility indicator to case incidence among 111 of the most populous U.S. counties during the Omicron surge from December 2021 through February 2022.

**Results:** Reported COVID-19 incidence peaked earlier and declined more rapidly among those counties exhibiting more extensive decline in mobility between December 20 and January 3. Based upon a fixed-effects, longitudinal cohort model, we estimated that every 1-percent decline in mobility between December 20 and January 3 was associated with a 0.63 percent decline in peak incidence during the week ending January 17 (95% confidence interval, 0.40-0.86 percent). Based upon a cross-sectional analysis including mean household size and vaccination participation as covariates, we estimated that the same 1-percent decline in mobility was associated with a 0.36 percent decline in cumulative reported COVID-19 incidence from January 10 through February 28 (95% CI, 0.18-0.54 percent).

**Conclusion:** Omicron did not simply sweep through the U.S. population until it ran out of susceptible individuals to infect. To the contrary, a significant fraction managed to avoid infection by engaging in risk-mitigating behaviors. More broadly, the behavioral response to perceived risk should be viewed as an intrinsic component of the natural course of epidemics in humans.

## Background

Prior to the emergence of the Omicron variant of SARS-CoV-2, numerous studies in various countries documented an association between a decline in population mobility and a subsequent reduction in reported case incidence [1-7]. The principal objective of the present study is to begin to assess whether this mobility-incidence relationship similarly prevailed during the more recent Omicron-driven wave.

There are several critical reasons why the mobility-incidence relationship observed for the ancestral strain and prior variants of SARS-CoV-2 may not apply equally to Omicron. More than any other variant, Omicron exhibited significant immune escape against vaccination and prior infection [8], though vaccines continued to protect against serious disease [9]. Omicron appears to have been about twice as transmissible as the Delta variant [10], with the larger proportion of asymptomatic Omicron infections likely enhancing the prevalence of super spreaders [11]. While home testing rose markedly in response to the initial news of the variant [12], later reports of Omicron’s tendency to spare the deep tissues of the lung [13] may have alleviated fears of serious illness that drive voluntary risk-mitigation behavior [14]. There is the further concern that frequently changing news reports and public health guidance induced “worry fatigue” [15], especially when perceptions of risk and compliance with such guidance are themselves subject to herd transmission [16, 17].

Mobility is a multidimensional concept that has been variously gauged by such diverse measures as smartphone visits to bars and restaurants [18], traffic patterns [7], and television watching as a proxy for time spent at home [19]. Here, following the lead of two key papers [20, 21], we employ the statistical technique of principal component analysis to collapse the six-dimensional Google Mobility Reports [22] into a single mobility indicator. Further adhering to a recent study of reported case incidence and hospitalization in relation to vaccination rates during the Delta surge [23], we restrict our analysis to the most populous counties in the United States, together comprising approximately 44 percent of the total U.S. population. Such an approach avoids the potential pitfalls of comparing small rural counties with large urban centers. We focus on the wave of reported cases from December 2021 through Feb 2022, during which Omicron was far and away the dominant variant.

## Methods

### Data: Most Populous Counties

We confined our analysis to the most populous counties in the United States. From an *initial sample* of all 112 counties with population exceeding 600,000, we excluded one county (Johnson County KS, population 602,000) as a result of missing data on one of the mobility measures to be described below. Our *analytic sample* thus consisted of 111 counties, together comprising 146.5 million persons or about 44 percent of the entire U.S. population. Supplement Fig. A maps the locations of all 112 counties in the initial sample, identifying the excluded county as well.

### Data: Google Mobility Reports

We relied upon Google Mobility Reports [22] to assess changes in mobility in each of the 111 counties in our analytic sample. Compiled from data on the movements of mobile devices, these reports provided daily measures of mobility for six distinct categories of places: retail & recreation; grocery & pharmacy; parks; transit stations; workplaces; and residential [24]. Based upon the number of visits to and length of stay in the places in each category, the reports showed activity as a percent of baseline, where the baseline represented the median value for the corresponding day of the week during the 5-week period from January 3 – February 6, 2020. For each of the 111 counties in the analytic sample and each of the six categories of mobility, we computed weekly mean values of mobility for the week ending Monday, February 24, 2020, through the week ending Monday, February 28, 2022. We chose a weekly ending date of Monday solely to be conformal with the available data on COVID-19 case reports, to be described below.

### Data: Community Profile Reports

We relied upon the COVID-19 Community Profile Reports, issued regularly by the White House COVID-19 Team [25], for data on the reported number of COVID-19 cases in each county for each week, starting with the week ending December 6, 2021, and continuing through the week ending February 28, 2022. We also relied upon this data source for estimates of each county’s population, from which we computed COVID-19 incidence rates, as well as two county-specific demographic characteristics: the U.S. Center for Disease Control’s social vulnerability index [26], and the average household size. We included the latter characteristic to capture the important influence of intra-household transmission on COVID-19 incidence [1].

### Data: County-Specific Vaccination

In addition to the foregoing county-specific demographic variables, we relied upon a database of COVID-19 vaccination participation rates, compiled by the U.S. Centers for Disease Control and Prevention [27]. These data showed the percentage of each county’s population who completed a one-or two-dose series of vaccinations, as well as the cumulative number of booster doses per 100 population, as of December 15, 2021, the earliest date for which both measures were available.

### Principal Component Analysis of Google Mobility Categories

We relied upon the data on the six weekly Google mobility measures in the 111-county database, covering the 106-week period from the week ending February 24, 2020, through the week ending February 28, 2022, to compute the first principal component as a summary measure of mobility [20, 21]. This summary measure, which we refer to here as our *mobility indicator*, represents the linear combination of the six individual mobility categories that captures the largest fraction of the overall variance of the data [28]. Denoting by *g*_*kit*_ the observed value of Google mobility category *k* in county *i* during week *t*, we thus computed the indicators 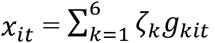, where the estimated coefficients *ζ*_*k*_ were not necessarily positive, but where 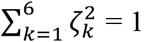.

### County-Specific Changes in Mobility

Having relied upon the entire database of multidimensional Google mobility categories from the week ending February 24, 2020, onward to compute our unidimensional mobility indicator, we then focused on the narrower 13-week period from the week ending December 6, 2021 through the week ending February 28, 2022, which encompassed the Omicron surge in the United States [29].

As described in detail in the Results below, we determined that our mobility indicator *x*_*it*_ (where *i* = 1, …, 111 and *t* = 1, …, 13) declined primarily during the interval between the week ending December 20, 2021 (that is, *t* = 3) to the week ending January 3, 2022 (that is, *t* = 5). For each county *i*, we thus computed the change in the mobility indicator Δ*x*_*i*_ = *x*_*i*5_ − *x*_*i*3_. Since mobility declined overall during the 13-week analysis period, the quantities Δ*x*_*i*_ were negative. We then divided the sample of counties into the lower half and upper half of the distribution of the absolute values |Δ*x*_*i*_|, denoting counties in the lower half as *less extensive mobility decline* and those in the upper half as *more extensive mobility decline*. We defined the binary variable *X*_*i*_ to equal 0 if county *i* was in the lower half of the distribution (less extensive decline) and 1 if county *i* was in the upper half of the distribution (more extensive decline).

### Longitudinal Cohort of Counties

The available data, described above, thus allowed us to construct a longitudinal cohort of 111 counties, indexed *i* = 1, …, 111, covering the 13-week period running from the week ending December 6, 2021 (*t* = 1) through the week ending February 28, 2022 (*t* = 13). For each county *i* and week *t*, we had data not only on our constructed mobility indicator *x*_*it*_, but also on *y*_*it*_, the incidence of reported cases of COVID-19 per 100,000 population.

To examine the qualitative relationships between changes in mobility and changes in COVID-19 incidence, we first plotted the population-weighted mean values of *x*_*it*_ and *y*_*it*_ over time for the two groups of counties with less extensive and more extensive declines in mobility. For example, the population-weighted mean mobility indicator among less-extensive-decline counties at week *t* would equal 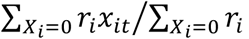, where *r*_*i*_ is the population of county *i* and where the summations are only over those counties *i* for which *X*_*i*_ = 0. The other conditional means were computed analogously.

To examine the quantitative relationships between changes in mobility and changes in COVID-19 incidence, we estimated a fixed-effects longitudinal cohort model with the following specification:

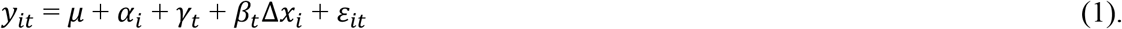

In equation (1), the parameter *μ* was an overall mean, while *α*_*i*_ and *γ*_*t*_ were county-specific and time-specific fixed effects, respectively. The parameters of interest *β*_*t*_ gauged the impact of county-specific changes in mobility on a week-by-week basis. Finally, *ε*_*it*_ were assumed to be spherical error terms. This fixed-effects model was estimated by ordinary least squares.

### Cross-Sectional Analyses

To further study the quantitative relationships between changes in mobility and changes in COVID-19 incidence, we defined the cumulative incidence for each county *i* during the period from week ending January 10, 2022 (*t* = 6) through the week ending February 28, 2022 (*t* = 13) as 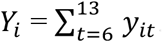. We then ran the cross-sectional model:

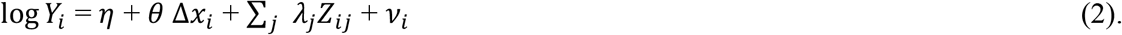

In equation (2), the parameter *η* was an overall mean, while the parameters *λ*_*j*_ captured the effects of county-specific covariates *Z*_*ij*_. The parameter of interest *θ* gauged the proportional impact the change in mobility during the period between December 20, 2021, and January 3, 2022, on the subsequent cumulative reported incidence of COVID-19 from the week ending January 10, 2022, onward. Finally, *ν*_*i*_ were assumed to be uncorrelated error terms. This cross-sectional model was estimated by population-weighted least squares.

### Test for Joint Causation

An alternative interpretation of the findings of models (1) and (2) was that *both* the change in mobility Δ*x*_*i*_ between December 20 and January 3 and the subsequent path of reported incidence *y*_*it*_ during January were jointly determined by the initial rate of acceleration of cases. If so, then inclusion of the initial acceleration rate in the model would attenuate any observed correlation between changes in mobility and subsequent changes in incidence. To address this possibility, we tested the following cross-sectional model:

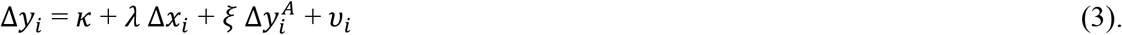

In equation (3), Δ*y*_*i*_ = *y*_*i*7_ − *y*_*i*6_ denotes the change in incidence between January 10 and 17, when cases were peaking, while 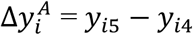 denotes the initial acceleration of incidence between December 27 and January 3. As in the previous models, *κ, λ*, and *ξ* were unknown parameters, while the *υ*_*i*_ were assumed to be uncorrelated error terms. As in model (2), equation (3) was estimated by population-weighted least squares. If the initial acceleration of COVID-19 cases in each county 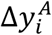 jointly determined both the mobility response Δ*x*_*i*_ and the subsequent path of reported incidence Δ*y*_*i*_, then inclusion of the term 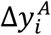 as an explanatory variable in equation (3) would result in an estimate of *λ* = 0.

## Results

### Mobility Indicator

Supplement Table A displays the estimated coefficients of the first principal component of the six Google mobility categories. The Google Retail & Recreation category of mobility had the largest contribution to the overall variance of our computed mobility indicator, while the Parks category had the smallest contribution. The Residential category had a negative estimated coefficient, inasmuch as increases in visits to and duration of stay in residences reflected a decrease in overall mobility.

Fig. 1 illustratively graphs the six Google mobility categories specifically for Philadelphia County, Pennsylvania (population 1,584,000), during the period from the week ending December 6, 2021 (*t* = 1), through the week ending January 31, 2022 (*t* = 9). Each of the colored piecewise linear plots shows the evolution of one of the original Google mobility categories *g*_*kit*_. The thicker black plot shows the corresponding evolution of our unidimensional mobility indicator *x*_*it*_, calculated from the coefficients in Supplement Table A. The path of this overall mobility indicator shows a significant decline during the two-week interval between the week ending December 20, 2021 (*t* = 3) and the week ending January 3, 2022 (*t* = 5).

**Fig. 1.**
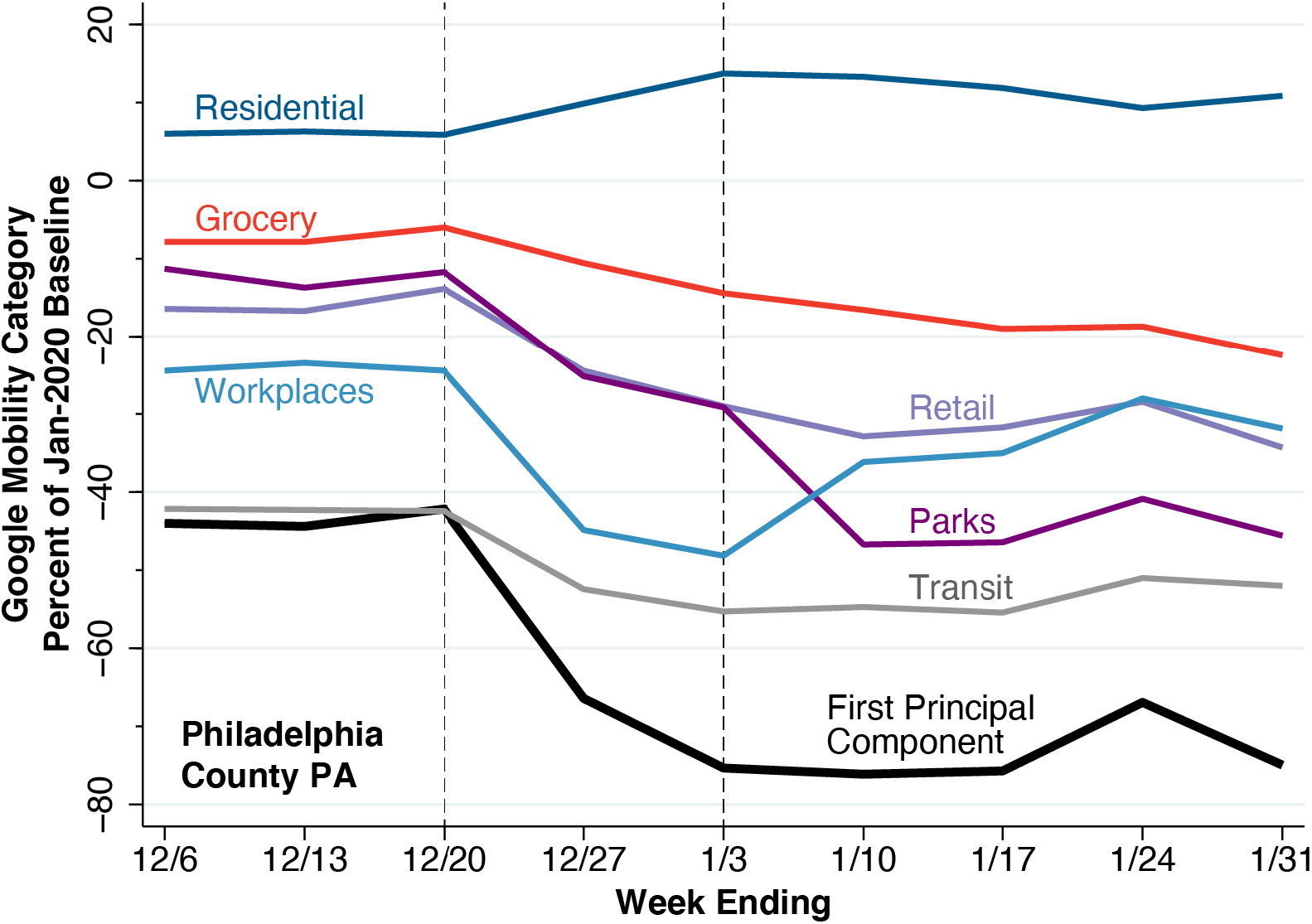
Construction of Mobility Indicator as First Principal Component of Six Google Mobility Categories, Philadelphia County, Pennsylvania, Weeks Ending December 6, 2021, Through January 31, 2022. Each colored piecewise linear plot shows the evolution of the weekly mean of one of the six Google mobility categories. The thicker, black piecewise linear plot shows the first principal component of the six mobility categories as calculated for Philadelphia County. The drop in the calculated mobility indicator occurred during the two-week interval from week ending 12/20/21 to the week ending 1/3/22.

Among the 111 counties under study, we observed a median absolute decline of 31.85 units in our unidimensional mobility indicator during the two-week interval between the week ending December 20, 2021 (*t* = 3) and the week ending January 3, 2022 (*t* = 5). Thus, we classified counties into the less-extensive-decline group (*X*_*i*_ = 0) if their observed absolute mobility decline was less than this median value and into the more-extensive-decline group (*X*_*i*_ = 1) if their absolute mobility decline was greater than this median value.

Fig. 2 displays illustrative paths of our unidimensional mobility indicator for 14 randomly selected counties in less-extensive-decline group (*X*_*i*_ = 0, left panel) and another 14 randomly selected counties in the more-extensive-decline group (*X*_*i*_ = 1, right panel). In both panels, we have highlighted the portions of each path covering the 2-week interval from December 10, 2021 – January 3, 2022. For the less-extensive-decline (*X*_*i*_ = 0) counties on the left, nearly all the calculated decline in overall mobility occurred during the first week. For the more-extensive-decline (*X*_*i*_ = 1) counties on the right, the calculated mobility indicators continued to decline during the second week.

**Fig. 2.**
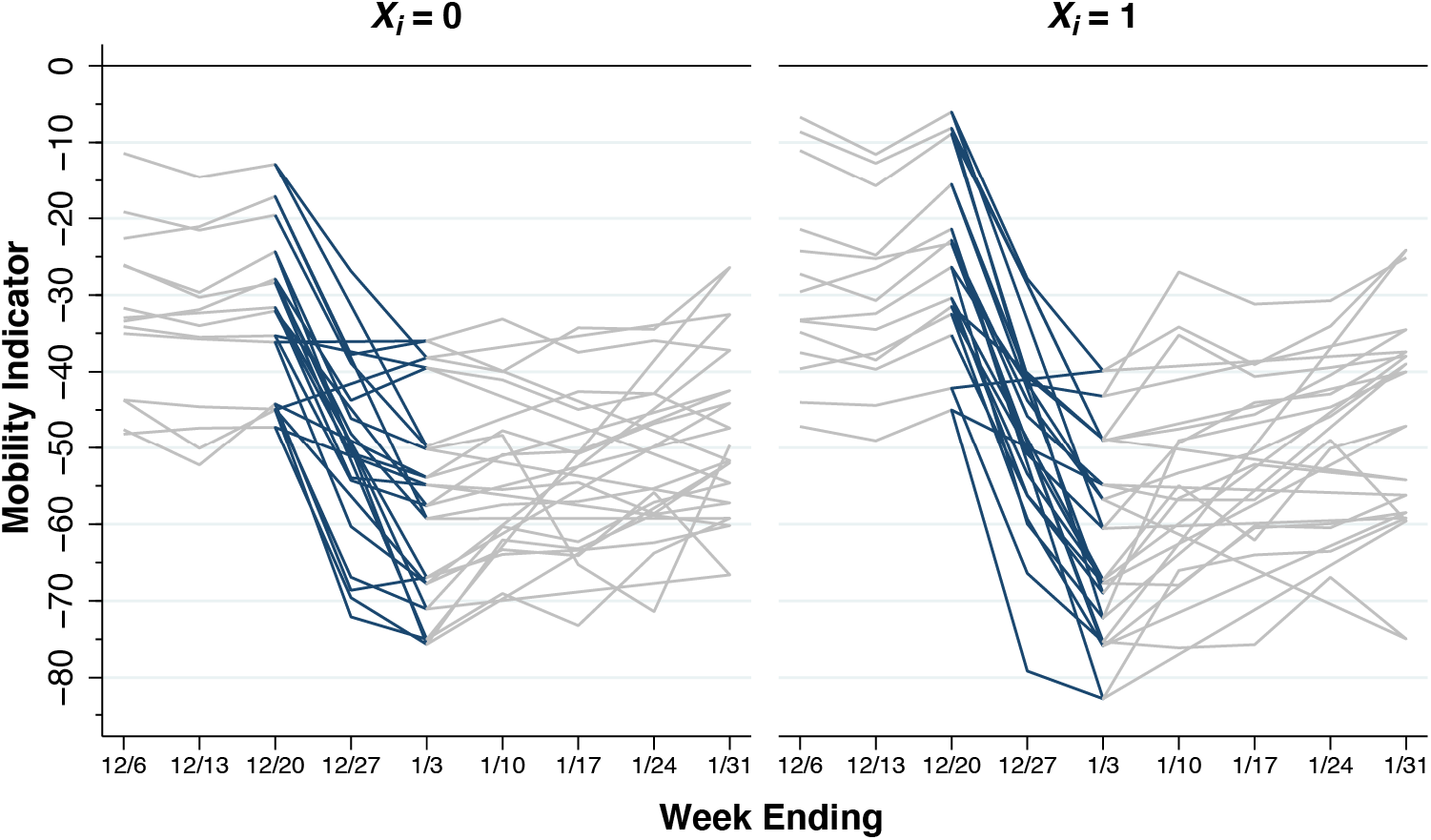
Illustrative Paths of Mobility indicator in Counties with Less Extensive and More Extensive Declines During the Two-Week Interval Between December 20, 2021, and January 3, 2022. The panel on the left, identified as *X*_*i*_ = 0, shows the paths of 14 randomly selected counties with an absolute decline of less than 31.85. The panel of the right, identified as *X*_*i*_ = 1, shows the paths of another 14 randomly selected counties with an absolute decline more than 31.85. In both panels, the paths of the calculated mobility indicators during the interval from 12/20/21 – 1/3/22 have been highlighted. For the less-extensive-decline (*X*_*i*_ = 0) counties on the left, nearly all the calculated decline in mobility occurred during the first week, that is, during 12/20 – 12/27/21. For the more-extensive-decline (*X*_*i*_ = 1) counties on the right, the calculated mobility indicators continued to decline during the second week, that is, during 12/27/21 – 1/3/22.

### Mobility and Case Incidence

Fig. 3 illustratively displays the combined paths of the mobility indicator *x*_*it*_ and the case incidence *y*_*it*_ specifically for Philadelphia County, Pennsylvania. The black-colored plot with square datapoints shows the path of the mobility indicator, replotted from the first principal component shown in Fig. 1, with the measurement scale along the left axis. The red-colored plot with circular datapoints, with measurement scale along the right axis, shows the path of COVID-19 case incidence in weekly reported cases per 100,000 population.

**Fig. 3.**
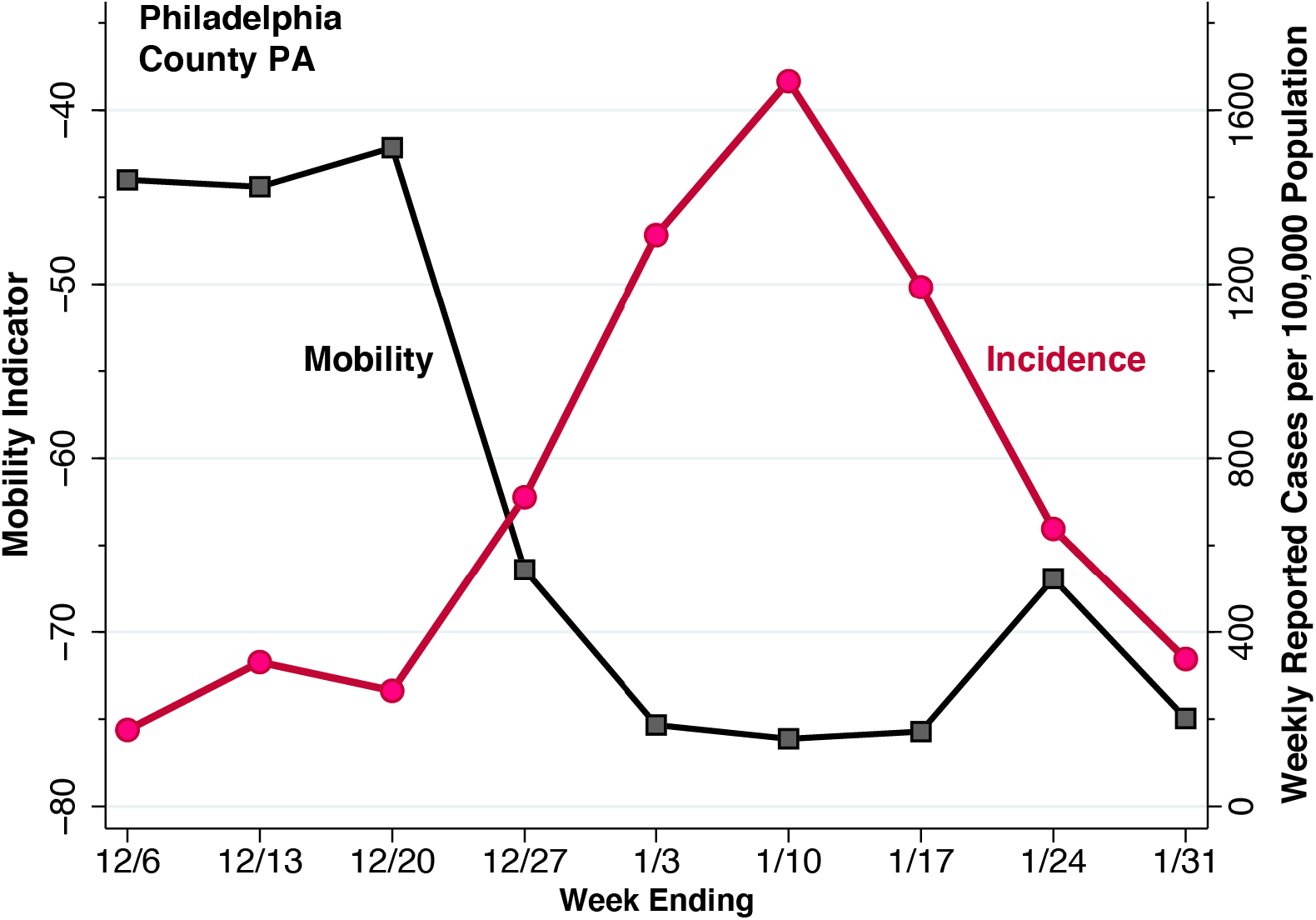
Mobility Indicator (Left Axis) and Reported COVID-19 Cases (Right Axis) in Philadelphia County, Pennsylvania, During the Weeks Ending December 6, 2021, Through January 31, 2022. The black plot with square datapoints corresponds to the weekly mobility indicator calculated in Fig. 1. The red plot with circular datapoints corresponds to the weekly counts of reported cases of COVID-19 per 100,000 population. Mobility declined during the interval from the week ending 12/20/21 through the week ending 1/3/22. The rise in reported cases peaked during the week ending 1/10/22 and declined thereafter.

For Philadelphia County PA, our computed mobility indicator *x*_*it*_ declined from –42.15 during the week ending December 20, 2021, to –75.33 during the week ending January 3, 2022. The observed absolute change of |Δ*x*_*i*_| = 33.18 thus placed Philadelphia County in the more-extensive-decline (*X*_*i*_ = 1) group. One week later, by the week ending January 10, 2022, reported COVID-19 incidence reached a peak of 1,666 cases per 100,000 population and declined thereafter.

### Longitudinal Cohort Analysis

For the less extensive and more extensive decline groups separately, Fig. 4 graphs the temporal paths of the population-weighted mean mobility indicator and population-weighted mean COVID-19 incidence during the 13-week study period. Both mobility and incidence have been computed as the change from the week ending December 6, 2021. Changes in mobility (square datapoints, measured on the left axis) are identified by the labels “Δ Mobility,” while changes in incidence (circular datapoints, right axis) are identified by the labels “Δ Incidence.”

**Fig. 4.**
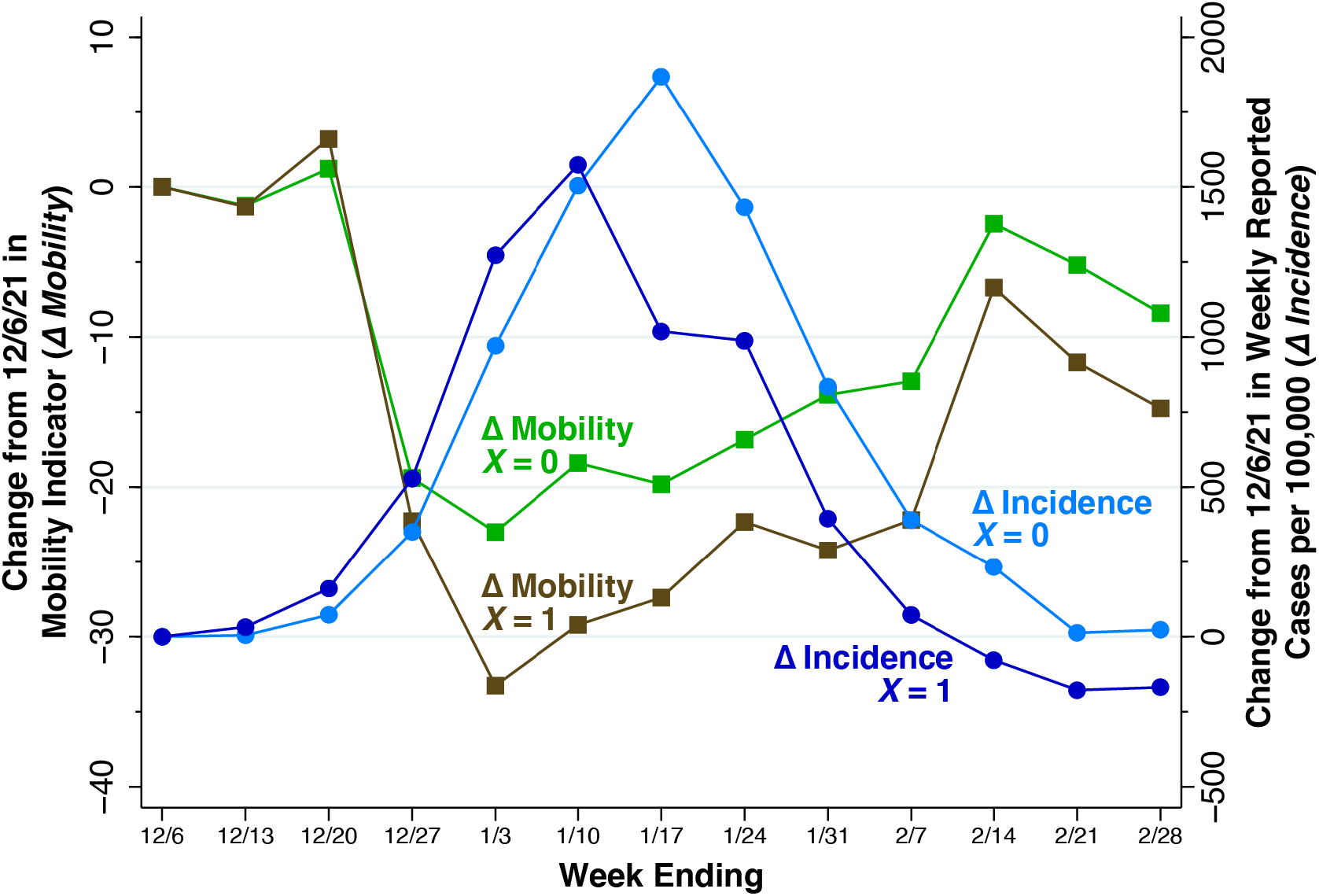
Changes in Mobility Indicators (Δ *Mobility*, Left Axis) and Changes in Reported COVID-19 Cases (Δ *Incidence*, Right Axis) in Less Extensive and More Extensive Mobility Decline Counties, Weeks Ending December 6, 2021, Through February 28, 2022. Less extensive mobility decline counties are identified as *X* = 0, while more extensive mobility decline counties are identified as *X* = 1. For both mobility and incidence, the figure plots the change from the week ending 12/6/21. Among less-extensive-decline (*X* = 0) counties, incremental incidence peaked during the week ending 1/17/22. Among more-extensive-decline (*X* = 1) counties, incremental incidence peaked earlier during the week of 1/10/22 and declined earlier.

Both the less extensive and more extensive decline counties followed essentially the same mobility path through the week ending December 27, 2021. During the subsequent week ending January 3, 2022, however, the two groups diverged, with the more-extensive-decline (*X* = 1) group exhibiting a larger continuing drop in mobility. These differences in mobility are reflected in the divergent paths of incremental COVID-19 incidence. Among less-extensive-decline (*X* = 0) counties, incidence peaked during the week ending January 17, while among more-extensive-decline (*X* = 1) counties, incidence reached a lower peak one week earlier.

Supplement Table B shows our estimates of the parameters of the fixed-effects model of equation (1). Fig. 5 below graphs the estimates of the key parameters of interest *β*_*t*_ for each week from *t* = 2, …, 13, as derived from that model. Since *t* = 1 (ending December 6, 2021) was the reference category, the parameter *β*_1_ was necessarily constrained to equal 1. The estimates of *β*_*t*_ from the week ending January 17 (*t* = 7) through the week ending February 14 (*t* = 11) are all positive and significant at the 5-percent level. For the peak week ending January 17, 2022, the estimated parameter was *β*_7_ = 37.7 with 95% confidence interval 23.9–51.4 (p < 0.001). That is, an additional one-point drop in our mobility indicator was associated with an incremental decline of 37.7 weekly reported cases of COVID-19 per 100,000 population.

**Fig. 5.**
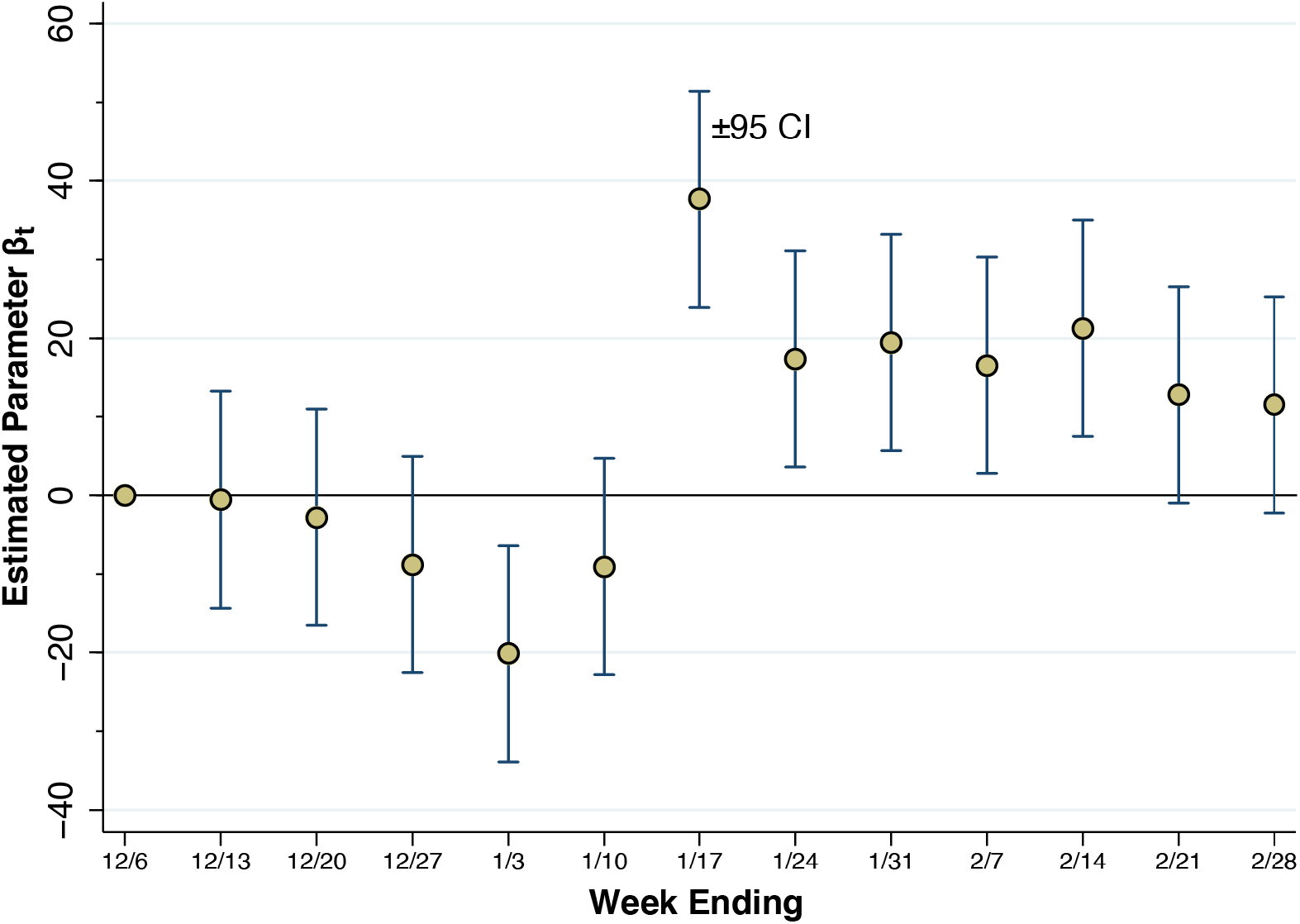
Estimates of the Interaction Parameters *β*_*t*_ in the Longitudinal Model of Equation (1). Except for the reference week ending 12/6/21, each week *t* has its own interaction parameter *β*_6_, which measures the marginal impact of a change in the mobility indicator from 12/20/21 to 1/3/22 (Δ *Mobility*) on the incremental reported incidence of COVID-19 cases per 100,000 population (Δ *Incidence*). The error bars surrounding each estimate are 95% confidence intervals. Starting with the week ending 1/17/22 and continuing through the week ending 2/14/22, the estimated parameters were positive and the null hypothesis of no association with changes in mobility could be rejected at the 5-percent level.

Based upon the observed sample means, we can reinterpret this estimated peak marginal effect *β*_7_ as an elasticity. With a population-weighted mean reported incidence of 1,729 cases per 100,000 during the week ending January 17, each one-point drop in mobility is thus associated with a 37.7/1729 = 2.18 percent drop in incidence (95% CI, 1.38–2.97 percent). At the population-weighted mean value of Δ*x* equal to –29.04, each one-point decrease represents a 1/29.04 = 3.44 percent decline in mobility. Thus, we obtain an estimated elasticity of 2.18/3.44 = 0.63 with a 95% confidence interval of 0.40–0.86.

### Cross-Sectional Results

Supplement Table C displays our estimates of the parameters of the cross-sectional model of equation (2). The estimated parameter *θ* was significantly different from zero in a bivariate specification on the change in mobility (specification A) as well as in multivariate specifications (B and C) that included demographic covariates and vaccination participation rates. Apart from the change in mobility, average household size was the only other explanatory variable exhibiting a statistically significant association with cumulative reported COVID-19 incidence.

Fig. 6 plots the cumulative cases per 100 population between the week ending January 10, 2022, and the week ending February 28, 2022 (that is, the variable *Y*_*i*_ in equation (2)) against the change in our calculated mobility indicator (Δ*x*_*i*_) between December 20, 2021, and January 3, 2022 (that is, the variable Δ*x*_*i*_). In accordance with the log-linear specification of equation (2), the vertical axis is measured on a logarithmic scale. The size of each datapoint reflects the county population.

**Fig. 6.**
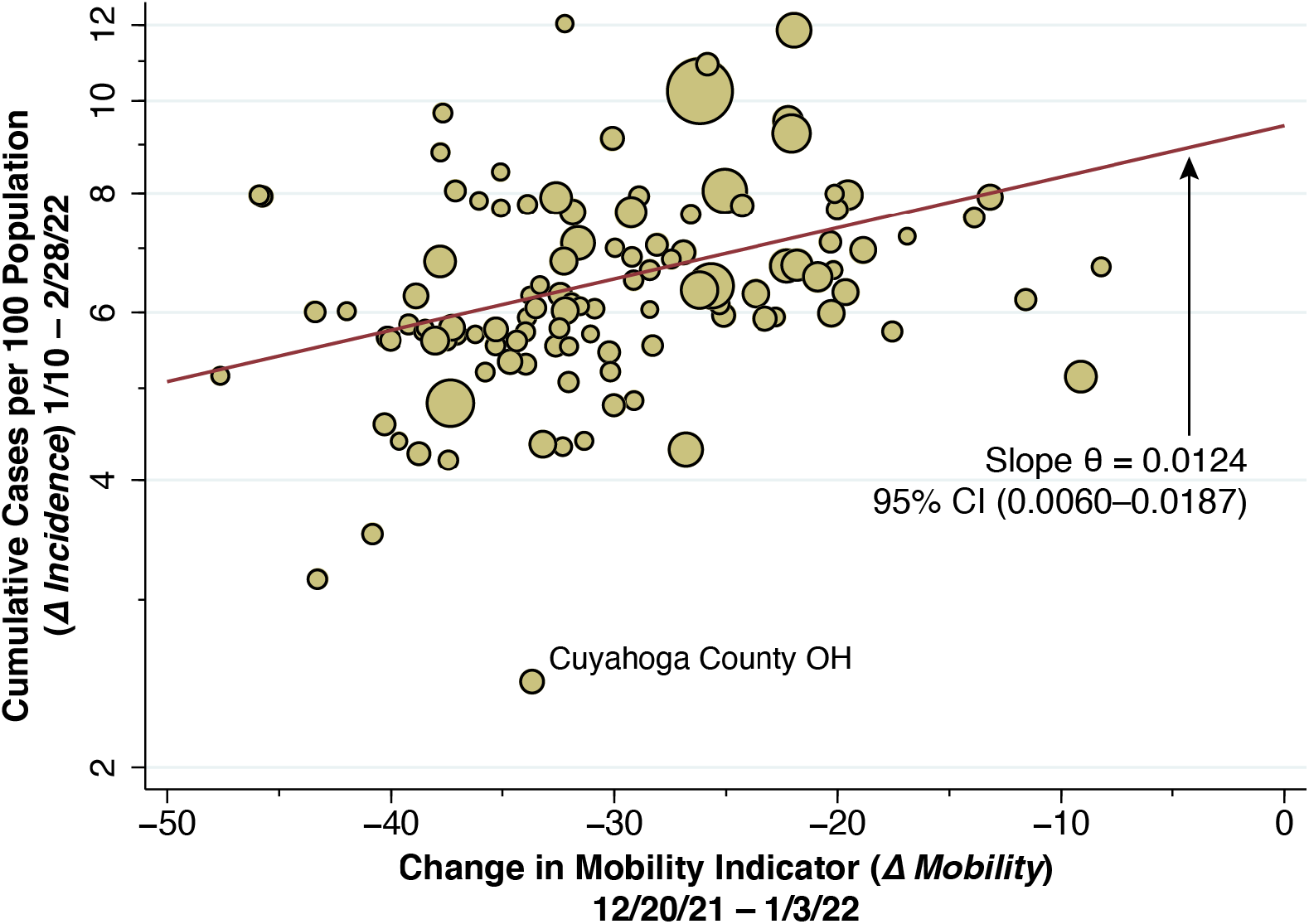
Cumulative Reported COVID-19 Cases per 100 Population During January 10 Through February 28, 2022 (Δ *Incidence*) Versus the Change in the Mobility Indicator During December 20, 2021, Through January 3, 2022 (Δ *Mobility*), Plotted for 111 Counties. The size of each datapoint reflects the county population. Cumulative reported COVID-19 cases are plotted on a logarithmic scale. The weighted least squares fitted line is shown in red. The estimated slope, corresponding to the parameter *θ* in equation (2) was 0.0124 with 95% CI (0.0060, 0.0187). That is, every additional 1-point reduction in the mobility indicator was associated with a 1.24 percent decline in cumulative reported cases per 100 persons. The outlier in the plot is identified as Cuyahoga County, Ohio. The vertical axis plots cumulative case incidence from the week ending January 10 onward. Cumulative incidence for the entire Omicron wave, from the week ending December 6, 2021, averaged 9 per 100 population.

The superimposed line represents the population-weighted least squares fit to the data. This corresponds to the bivariate regression of log *Y*_*i*_ versus Δ*x*_*i*_ without additional covariates *Z*_*i*_, shown as specification A in Supplement Table C. The estimate of the slope parameter *θ* was 0.0124, with 95% confidence interval 0.0060–0.0.187. We can similarly reinterpret this cumulative marginal effect as an elasticity. Thus, every additional one-point decrease in our calculated mobility indicator was associated with a 1.24 percent decline in cumulative case incidence. At the population-weighted mean value of Δ*x* equal to –29.04, each one-point decrease represents a 1/29.04 = 3.44 percent decline in mobility. Thus, we obtain an estimated elasticity of 1.24/3.44 = 0.36 with a 95% confidence interval of 0.18–0.54.

### Test of Joint Causation

Supplement Table D displays the estimates of our joint causation model (3). We found that the estimated coefficient *ξ* of the initial acceleration of reported incidence 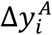 was negative. That is, early acceleration of COVID-19 incidence during the week after December 27 was associated with a decline from peak incidence during the week after January 10. However, inclusion of the covariate 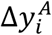 in the cross-sectional model did not materially affect the significant positive coefficient *λ* of the change in mobility Δ*x*_*i*_. Supplement Fig. B further shows graphically how the inclusion of the additional covariate 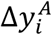 resulted in no material change in the fitted linear model relating the change in incidence Δ*y*_*i*_ between January 10 and January 17 to the change in mobility Δ*x*_*i*_ between December 20 and January 3.

## Discussion

### Interpreting Elasticities

For each one-percent decline in our unidimensional measure of mobility, we have estimated a 0.63-percent decline in peak reported case incidence (95% confidence interval, 0.40 to 0.86 percent) and a 0.36-percent decline in cumulative reported case incidence (95% confidence interval, 0.18 to 0.54 percent). That the short-term elasticity of peak incidence turns out to be greater than the longer-term elasticity of cumulative incidence is hardly unexpected. As the prevalence of infection falls beyond the peak of epidemic wave, the effectiveness of risk-avoidance measures would be expected to decline. The declining marginal effects derived from the longitudinal cohort model, as seen in Fig. 5, are consistent with this interpretation.

Nor is it unexpected that the estimated mobility-incidence elasticity should be less than 1 even at the peak of the Omicron wave. For it implies that there were some sources of infection whose risks could not be mitigated through the available mobility-reduction strategies. Consider, for example, an individual whose only source of infection was taking public transport to work. If she cut back her exposure through this modality by *x* percent, her risk of infection would likewise decline by *x* percent, and the mobility-incidence elasticity would be 1. If, on the other hand, intrahousehold transmission from family members was a second, independent source of infection, then her cutting back on public transport by *x* percent would lower her infection risk by less than *x* percent, and the corresponding elasticity would be less than unity. Our finding that average household size was a significant determinant of county-specific Omicron case incidence (Supplement Table C) suggests that this example is more than hypothetical.

### Change in Behavior as an Intrinsic Feature of Course of Epidemics in Humans

Our results belie the hypothesis that Omicron simply swept through the population until the variant ran out of susceptible individuals to infect. For the entire Omicron surge, cumulative reported incidence averaged approximately 9 cases per 100 population (Fig. 6). If only one-fourth of all Omicron infections were reported by public authorities [30], then approximately 36 percent of the population became infected during the Omicron surge. In view of Omicron’s documented capacity for immune escape from vaccination and prior infection [8], there had to be no small fraction of susceptible individuals who, by engaging in risk-mitigating behaviors, managed to avoid infection.

Our findings reinforce the broader conclusion that the behavioral response to perceived risk needs to be regarded as an intrinsic component of the course of epidemics in humans. Quite apart from the evidence now accumulated in the ongoing COVID-19 pandemic, such behavioral responses have been documented for HIV in developing countries [31], the SARS outbreak in Hong Kong [32], the swine flu outbreak [33], the H1N1 influenza outbreak [34], and sexually transmitted diseases generally [35].

### The Decline in Mobility Began Before the Peak in Disease Incidence

Theoretical treatments of the human behavioral response during an epidemic have generally adopted the ad hoc strategy of making the contact frequency between susceptible and infected persons an inverse function of the contemporary prevalence of infection [36-39]. The difficulty with this approach is that, as shown in Figs. 2–4, the decline in mobility occurred 2-3 weeks *before* the peak in reported incidence. One possible explanation is that changes in behavior were a response to extensive news about the upcoming surge in infections, rather than the surge itself.

Omicron emerged on the world scene in late November 2021 essentially as an unanticipated shock. The initial reaction to this shock was a wave of news reports through the first three weeks of December, bracing the country for the coming surge of cases and hospitalizations [40-45]. According to Google Trends data for the U.S. [46, 47], searches for “omicron” initially rose at the end of November and then surged during the third week of December, reaching a peak on December 21, while searches for “covid omicron symptoms” subsequently peaked on December 27, 2021. Robust models of changes in behavior during an epidemic need to account for the critical intervening role of the media [48-51].

### The Dynamics of a Natural Experiment

Our findings can be thus interpreted as the result of a natural experiment precipitated by the unanticipated shock of Omicron’s emergence. The widespread decline in mobility across multiple counties, observed in Fig. 2, was a reaction to the rapid, nationwide diffusion of the news about the new variant. While these mobility reactions were closely aligned temporally, their magnitudes varied nontrivially. As a result of these geographic variations in the extent of mobility decline, we observed subsequent variations in the depth of the variant’s penetration across communities. Thus, an initial shock across an entire country produced responses of variable magnitude intended to modulate the shock, which in turn led to dynamic variations in the ultimate impact of the shock.

### The Argument for Reverse Causation

The principal objection to this interpretation is that the near-coincident declines in mobility were not random and, accordingly, our study design cannot demonstrate a purely causal relation between mobility and the incidence of infection. To the contrary, the argument goes, the observed declines in our unidimensional mobility indicator between the week ending December 10, 2021, and the week ending January 3, 2022, could also have been an early *response* to the emerging Omicron wave. One might conjecture, in fact, that the somewhat greater COVID-19 incidence in high-mobility-decline counties seen in Fig. 4, especially during the week ending January 3, was in fact the stimulus for the inhabitants of those counties to continue to engage in mobility-reducing behaviors. Such an interpretation would seem to square with the significant negative estimate of the parameter *β*_5_ in Fig. 5.

In view of such reverse causation, our estimates of the parameters *β*_7_ through *β*_13_ in Fig. 5, covering the period from the week ending January 13 onward, may indeed be biased upward, as is our cross-sectional slope parameter *θ* in Fig. 6. However, the results of our joint causation model (Supplement Table D and Fig. B) suggest that the magnitude of this bias is likely to be small. In short, the striking temporal relation between the extent of the mobility reductions observed through the week ending January 3 and the *subsequent* divergence in COVID-19 incidence, as seen in Fig. 4, cannot readily be explained by reverse causation.

### Policy Endogeneity

It would have been preferable, some might contend, to instead construct predictor variables based upon the extent of policy restrictions on mobility imposed in each county, such as renewed requirements on indoor mask use. In principle, such restrictions would be regarded as exogenous instruments to identify the unbiased effect of the endogenous mobility indicator that we have relied upon here [52]. The problem with this approach is that policies intended to restrict mobility are likewise endogenous. A public authority’s decision to impose a mask mandate may just as well be a response to news of rising COVID-19 cases as an individual’s uncoerced decision not to take the subway.

There is little basis to suppose, in any event, that declines in mobility such as those consistently observed in Figs. 1 through 4 are necessarily responses to coercive measures by public authorities. The near collapse of subway ridership in New York City during the second week of March 2020 was followed within 1–2 weeks by the flattening of the COVID-19 incidence curve. Yet no government authority ordered New Yorkers to stop taking the subway en masse [2].

### Appropriateness of a Unidimensional Mobility Indicator

The data in Figs. 1 through 4 make a strong case in favor of the suitability of our unidimensional summary indicator of the six Google mobility categories. In the illustrative plot in Fig. 1, we saw how five of the individual categories tended to move together, while the residential category tended to move in the opposite direction. Our principal component analysis (Supplement Table A) confirmed these observations and further demonstrated that visits to retail establishments captured a larger fraction of the overall variance of the six categories. In the illustrative plot of Fig. 2, we saw how the resulting unidimensional indicator consistently captured changes in mobility during the two-week interval from the week ending December 20 to the week ending January 3. In Figs. 3 and 4, we saw how the temporal path of our unidimensional mobility indicator during that interval was followed by a peaking in reported Omicron cases 2-3 weeks later.

### Cross-Sectional Analysis with Covariates

In our longitudinal cohort analysis of equation (1), we relied on the statistical technique of fixed effects to capture other, persistent unobserved characteristics of individual counties. In the cross-sectional analysis of equation (2), by contrast, we relied upon county-specific demographic variables and indicators of vaccination participation. Unfortunately, we did not have county-specific data on booster vaccinations before December 15, 2021. Consequently, our data may include a nontrivial number of recent vaccinations in response to emerging news about the coming Omicron wave.

In contrast to our longitudinal study of a cohort of 111 counties over 13 successive weeks, our cross-sectional analysis encompassed only 111 county-specific observations on cumulative reported COVID-19 incidence. As already noted, reported cases of Omicron may have constituted no more than one-quarter of all incident cases [30]. This observation raises the possibility that the degree of underreporting in a particular county was related to the magnitude of the observed decline in mobility. To the extent that counties with a higher perceived risk and greater self-imposed declines in mobility also reported more cases, our cross-sectional estimates would understate the strength of the mobility-incidence relationship.

## Conclusion

This study documented a striking dynamic relationship between declines in mobility and subsequently reported reductions in case incidence during the Omicron surge in the most populous counties in the United States. The mobility-incidence relation prevailed despite the high degree of immune escape by the Omicron variant, as well as the potentially dissuasive effects of so-called worry fatigue on risk-mitigating behavior. Our findings imply that a significant fraction of the population managed to avoid infection by engaging in risk-mitigating behaviors. More broadly, the behavioral response to perceived risk should be viewed as an intrinsic component of the natural course of epidemics in humans.

## Data Availability

Supporting programs and data have been posted at https://osf.io/6edyc/.

## Declarations

### Availability of Data and Materials

Supporting programs and data have been posted on the Open Science Framework at https://osf.io/6edyc/.

## Abbreviations

None.

## Acknowledgments

This article represents the sole opinion of its author and does not necessarily represent the opinions of the Massachusetts Institute of Technology, Eisner Health, or any other organization.

## Human Subjects and Confidentiality

This study relies exclusively on publicly available data that contain no individual identifiers.

## Prior Publication

An earlier version of this study has been posted on the medRxiv.org preprint server at https://www.medrxiv.org/content/10.1101/2022.03.16.22272523v1.

