## Supplement for "Mobility was a Significant Determinant of Reported COVID-19 Incidence During the Omicron Surge in the Most Populous U.S. Counties"

### Supplement: Mobility and COVID-19 Incidence During the Omicron Surge

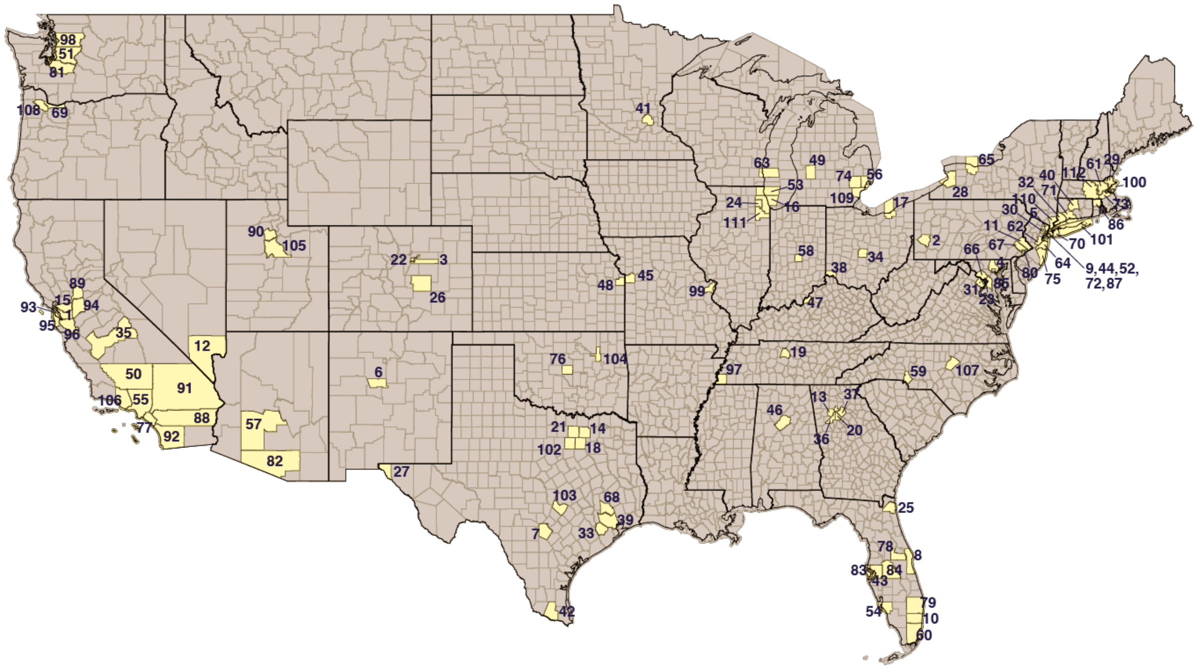


**Fig. A. Initial Sample of 112 U.S. Counties with Population ≥ 600,000.** All counties are numbered in accordance with the legend below. State boundaries are indicated by the thicker black lines. Johnson County KS (45) was excluded due to missing data on transit mobility.

| # | County | State | Pop (000) | # | County | State | Pop (000) | # | County | State | Pop (000) | # | County | State | Pop (000) |
|---|---|---|---|---|---|---|---|---|---|---|---|---|---|---|---|
| 1 | Alameda | CA | 1,671 | 29 | Essex | MA | 789 | 57 | Maricopa | AZ | 4,485 | 85 | Prince George's | MD | 909 |
| 2 | Allegheny | PA | 1,216 | 30 | Essex | NJ | 799 | 58 | Marion | IN | 965 | 86 | Providence | RI | 639 |
| 3 | Arapahoe | CO | 657 | 31 | Fairfax | VA | 1,148 | 59 | Mecklenburg | NC | 1,110 | 87 | Queens | NY | 2,254 |
| 4 | Baltimore | MD | 827 | 32 | Fairfield | CT | 943 | 60 | Miami-Dade | FL | 2,717 | 88 | Riverside | CA | 2,471 |
| 5 | Bergen | NJ | 932 | 33 | Fort Bend | TX | 812 | 61 | Middlesex | MA | 1,612 | 89 | Sacramento | CA | 1,552 |
| 6 | Bernalillo | NM | 679 | 34 | Franklin | OH | 1,317 | 62 | Middlesex | NJ | 825 | 90 | Salt Lake | UT | 1,160 |
| 7 | Bexar | TX | 2,004 | 35 | Fresno | CA | 999 | 63 | Milwaukee | WI | 946 | 91 | San Bernardino | CA | 2,180 |
| 8 | Brevard | FL | 602 | 36 | Fulton | GA | 1,064 | 64 | Monmouth | NJ | 619 | 92 | San Diego | CA | 3,338 |
| 9 | Bronx | NY | 1,418 | 37 | Gwinnett | GA | 936 | 65 | Monroe | NY | 742 | 93 | San Francisco | CA | 882 |
| 10 | Broward | FL | 1,953 | 38 | Hamilton | OH | 817 | 66 | Montgomery | MD | 1,051 | 94 | San Joaquin | CA | 762 |
| 11 | Bucks | PA | 628 | 39 | Harris | TX | 4,713 | 67 | Montgomery | PA | 831 | 95 | San Mateo | CA | 767 |
| 12 | Clark | NV | 2,267 | 40 | Hartford | CT | 892 | 68 | Montgomery | TX | 607 | 96 | Santa Clara | CA | 1,928 |
| 13 | Cobb | GA | 760 | 41 | Hennepin | MN | 1,266 | 69 | Multnomah | OR | 813 | 97 | Shelby | TN | 937 |
| 14 | Collin | TX | 1,035 | 42 | Hidalgo | TX | 869 | 70 | Nassau | NY | 1,357 | 98 | Snohomish | WA | 822 |
| 15 | Contra Costa | CA | 1,154 | 43 | Hillsborough | FL | 1,472 | 71 | New Haven | CT | 855 | 99 | St. Louis | MO | 994 |
| 16 | Cook | IL | 5,150 | 44 | Hudson | NJ | 672 | 72 | New York | NY | 1,629 | 100 | Suffolk | MA | 804 |
| 17 | Cuyahoga | OH | 1,235 | 45 | Jackson | MO | 703 | 73 | Norfolk | MA | 707 | 101 | Suffolk | NY | 1,477 |
| 18 | Dallas | TX | 2,636 | 46 | Jefferson | AL | 659 | 74 | Oakland | MI | 1,258 | 102 | Tarrant | TX | 2,103 |
| 19 | Davidson | TN | 694 | 47 | Jefferson | KY | 767 | 75 | Ocean | NJ | 607 | 103 | Travis | TX | 1,274 |
| 20 | DeKalb | GA | 759 | 48 | Johnson* | KS | 602 | 76 | Oklahoma | OK | 797 | 104 | Tulsa | OK | 652 |
| 21 | Denton | TX | 887 | 49 | Kent | MI | 657 | 77 | Orange | CA | 3,176 | 105 | Utah | UT | 636 |
| 22 | Denver | CO | 727 | 50 | Kern | CA | 900 | 78 | Orange | FL | 1,393 | 106 | Ventura | CA | 846 |
| 23 | Dist. Columbia | DC | 706 | 51 | King | WA | 2,253 | 79 | Palm Beach | FL | 1,497 | 107 | Wake | NC | 1,112 |
| 24 | DuPage | IL | 923 | 52 | Kings | NY | 2,560 | 80 | Philadelphia | PA | 1,584 | 108 | Washington | OR | 602 |
| 25 | Duval | FL | 958 | 53 | Lake | IL | 697 | 81 | Pierce | WA | 905 | 109 | Wayne | MI | 1,749 |
| 26 | El Paso | CO | 720 | 54 | Lee | FL | 771 | 82 | Pima | AZ | 1,047 | 110 | Westchester | NY | 968 |
| 27 | El Paso | TX | 839 | 55 | Los Angeles | CA | 10,039 | 83 | Pinellas | FL | 975 | 111 | Will | IL | 691 |
| 28 | Erie | NY | 919 | 56 | Macomb | MI | 874 | 84 | Polk | FL | 725 | 112 | Worcester | MA | 831 |

Table A below displays the estimated coefficients ($\zeta_k$, $k$ = 1, …, 6) derived from the principal component analysis of Google mobility measures. The Residential measure has a negative coefficient, as increases in visits and duration of stay at residences reflect a decrease in mobility. Aside from the Parks mobility measure, all coefficients have approximately the same absolute values.

| **Table A. Estimated Coefficients and Unexplained Variances of the First Principal Component of Six Google Mobility Measures** | | |
|---|---|---|
| *Google Mobility Measure* | *Coefficient* | *Proportion Unexplained Variance* |
| Retail & Recreation | 0.4701 | 0.095 |
| Grocery & Pharmacy | 0.4188 | 0.282 |
| Parks | 0.2037 | 0.830 |
| Transit Stations | 0.4183 | 0.284 |
| Workplaces | 0.4223 | 0.270 |
| Residential | –0.4571 | 0.145 |

Table B shows the estimates of the parameters ($\beta_t$, $t$ = 2, …, 13) of the fixed-effects model of equation (1). Parameter $\beta_1$ for the reference category was set equal to 1.

| **Table B. Estimated Parameters of the Fixed-Effects Model of Equation (1)*** | | |
|---|---|---|
| *Parameter* | *Estimate* | *95% Confidence Interval* |
| $\beta_2$ | –0.549 | –14.315, 13.247 |
| $\beta_3$ | –2.816 | –16.582, 10.950 |
| $\beta_4$ | –8.810 | –22.575, 4.957 |
| $\beta_5$ | –20.146 | –33.913, –6.380 |
| $\beta_6$ | –9.069 | –22.835, 4.607 |
| $\beta_7$ | 37.663 | 23.896, 51.429 |
| $\beta_8$ | 17.331 | 3.573, 31.106 |
| $\beta_9$ | 19.421 | 5.655, 33.188 |
| $\beta_{10}$ | 16.537 | 2.771, 30.304 |
| $\beta_{11}$ | 21.235 | 7.469, 35.001 |
| $\beta_{12}$ | 12.777 | –0.989, 26.543 |
| $\beta_{13}$ | 11.500 | –2.266, 25.266 |
| *Total of 1,443 observations on 111 groups (13 observations per group). $R^2$= 0.673. Model estimated by ordinary least squares. | | |

Table C below shows the results of our cross-section model of equation (2). Specification A includes only the county specific change in mobility ($\Delta x$) as a covariate. Specification B adds average household size as a covariate, while specification C includes the social vulnerability index and our two indicators of vaccination participation.

| **Table C. Estimated Parameters of the Cross-Sectional Model of Equation (2).*** | | | |
|---|---|---|---|
| *Explanatory Variable* | *Specification A* | *Specification B* | *Specification C* |
| Change in Mobility Indicator ($\Delta x$) | **0.0124** (0.0032) | **0.0096** (0.0032) | **0.0090** (0.0033) |
| Average Household Size | | **0.2830** (0.0904) | **0.2524** (0.0972) |
| Social Vulnerability Index | | | 0.1643 (0.1172) |
| Percent Vaccine Series Complete | | | 0.0048 (0.0030) |
| Booster Doses per 100 Population | | | –0.0007 (0.0038) |
| Constant Term | **2.2435** (0.0959) | **1.3836** (0.2897) | **1.0638** (0.3405) |
| $R^2$ statistic | 0.1205 | 0.1937 | 0.2243 |

*In all specifications, the dependent variable was the logarithm of the cumulative case incidence per 100 population between the week ending January 10, 2022 ($t$ = 6) and the week ending February 28, 2022 ($t$ = 13) per 100 population. There was a total of 111 observations. All models estimated by population-weighted least squares. Highlighted in bold are parameter estimates significantly different from 0 (2-sided t-test, p < 0.05). Standard errors are shown in parentheses below each parameter estimate.

Table D below shows estimates of the parameters of joint causation model (3). In Specification A, which tests the bivariate relation between $\Delta y_i$ and the change in mobility indicator $\Delta x_i$, the estimated parameter $\lambda$ was positive and significantly different from zero (2-sided t-test, p < 0.001). In Specification B, which tests the bivariate relation between $\Delta y_i$ and the initial acceleration of cases $\Delta y_i^A$, the estimated parameter $\xi$ was negative and significantly different from zero (2-sided t-test, p = 0.001). That is, early acceleration of the Omicron wave in a particular county was associated with subsequent early reversal of reported COVID-19 incidence by January 10, 2022 ($t$ = 7). However, in Specification B, inclusion of both mobility indicator $\Delta x_i$ and the initial acceleration of cases $\Delta y_i^A$ had a minimal effect on the estimated parameter $\lambda$, which remained positive and significantly different from zero (2-sided t-test, p = 0.001). Moreover, the hypothesis that $\lambda$ was no different from its estimated value in Specification A could not be rejected (2-sided F-test, p = 0.534).

| Table D. Estimated Parameters of the Joint Causation Model of Equation (3).* | | | |
|---|---|---|---|
| *Explanatory Variable* | *Specification A* | *Specification B* | *Specification C* |
| Change in Mobility Indicator ($\Delta x$) | **57.650**<br>(9.769) | | **51.005**<br>(9.230) |
| Acceleration of Incidence ($\Delta y^A$) | | **–0.6454**<br>(0.1452) | **–0.5348**<br>(0.1298) |
| Constant Term | **1675.274**<br>(293.168) | **432.321**<br>(124.431) | **1865.563**<br>(277.660) |
| $R^2$ statistic | 0.2421 | 0.1535 | 0.2243 |

*In all specifications, the dependent variable was the change in reported COVID-19 incidence per 100 population between the week ending January 10, 2022 ($t$ = 6) and the week ending January 17, 2022 ($t$ = 7). There was a total of 111 observations. All models estimated by population-weighted least squares. Highlighted in bold are parameter estimates significantly different from 0 (2-sided t-test, p < 0.05). Standard errors are shown in parentheses below each parameter estimate.

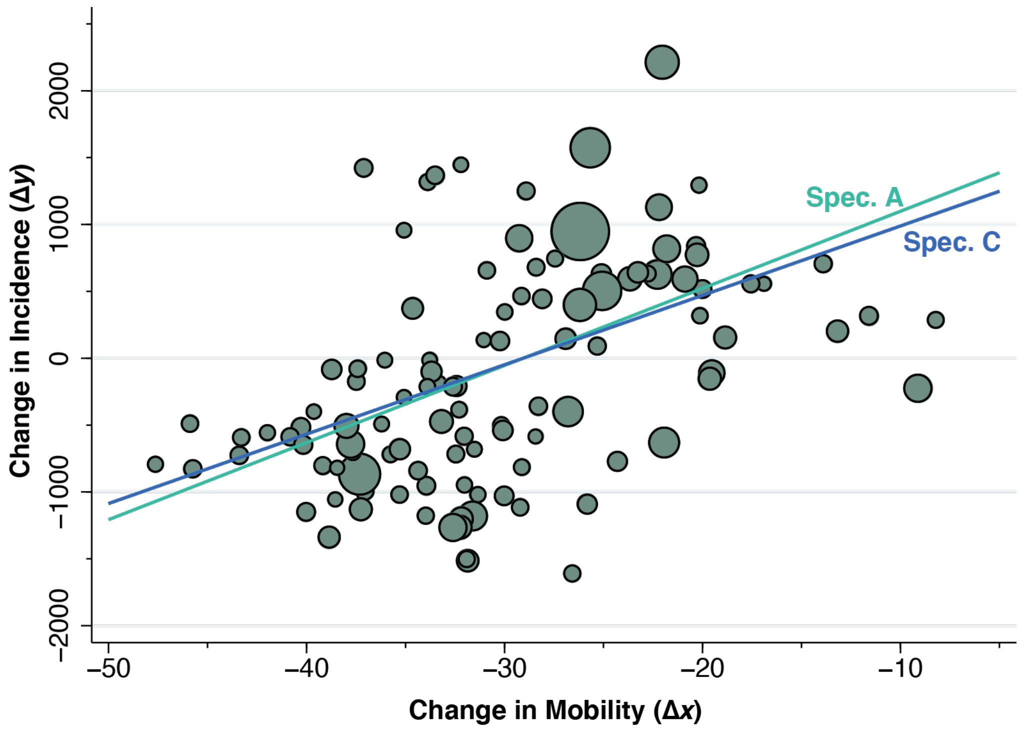


**Fig. B. Change in Reported Incidence Between January 10 and January 17 ($\Delta y$) Versus Change in Mobility Between December 20 and January 3 ($\Delta x$).** The size of each point reflects the county population. The line identified as Spec. A shows the population-weighted least squares fit to the data from Specification A in Table D. The line identified as Spec. C shows the corresponding fit from Specification C under the assumption at the initial acceleration ($\Delta y^A$) took on its population-weighted mean value. While Specification C resulted in a slightly lower slope, the null hypothesis of equal slopes could not be rejected (p = 0.534).

Fig. B shows a scatterplot underlying the results of Table D. The vertical axis measures $\Delta y_i$, that is, the change in reported COVID-19 incidence per 100 population between the week ending January 10, 2022 ($t$ = 6) and the week ending January 17, 2022 ($t$ = 7). The horizontal axis measures $\Delta x_i$, that is, the change in mobility between December 20 and January 3. The size of each data point reflects the county population size.

The two fitted lines represent the results of Specifications A and C. The line for Specification A plots the equation $\kappa + \lambda\, \Delta x_i$, based upon the estimated values of $\kappa$ and $\lambda$. The line for Specification C plots the equation $\kappa + \lambda\, \Delta x_i + \xi\, \overline{\Delta y^A}$, based upon the estimated values of $\kappa$, $\lambda$, and $\xi$, where $\overline{\Delta y^A}$ represents the population-weighted mean value of the initial acceleration $\Delta y_i^A$ across counties. We can see that the two estimated fitted lines are nearly indistinguishable, and in fact, the null hypothesis of equal slopes could not be rejected (p = 0.534).
